# Validation of an unbiased metagenomic detection assay for RNA viruses in viral transport media and plasma

**DOI:** 10.1101/2024.03.26.24304688

**Authors:** Anthony D. Kappell, Kathleen Q. Schulte, Elizabeth A. Scheuermann, Matthew B. Scholz, Nicolette C. Keplinger, Amanda N. Scholes, Taylor A. Wolt, Viviana M. June, Cole J. Schulte, Leah W. Allen, Krista L. Ternus, F. Curtis Hewitt

## Abstract

Unbiased long read sequencing holds enormous potential for the detection of pathogen sequences in clinical samples. However, the untargeted nature of these methods precludes conventional PCR approaches, and the metagenomic content of each sample increases the challenge of bioinformatic analysis. Here, we evaluate a previously described novel workflow for unbiased RNA virus sequence identification in a series of contrived and real-world samples. The novel multiplex library preparation workflow was developed for the Oxford Nanopore Technologies (ONT) MinION^TM^ sequencer using reverse transcription, whole genome amplification, and ONT’s Ligation Sequencing Kit with Native Barcode Expansion. The workflow includes spiked MS2 Phage as an internal positive control and generates an 8-plex library with 6 samples, a negative control and a *gfp* transcript positive control. Targeted and untargeted data analysis was performed using the EPI2ME Labs framework and open access tools that are readily accessible to most clinical laboratories. Contrived samples composed of common respiratory pathogens (Influenza A, Respiratory Syncytial Virus and Human Coronavirus 229E) in viral transport media (VTM) and bloodborne pathogens (Zika Virus, Hepatitis A Virus, Yellow Fever Virus and Chikungunya Virus) in human plasma were used to establish the limits of detection for this assay. We also evaluated the diagnostic accuracy of the assay using remnant clinical samples and found that it showed 100% specificity and 62.9% clinical sensitivity. More studies are needed to further evaluate pathogen detection and better position thresholds for detection and non-detection in various clinical sample metagenomic mixtures.

## Introduction

RNA viruses pose a significant threat to global public health. The prime example of this is the COVID-19 pandemic, but it is far from the only virus that has caused outbreaks in recent years. There are seasonal outbreaks every year of respiratory RNA viruses, such as paramyxoviruses (e.g., respiratory syncytial virus and human metapneumovirus) and alphainfluenzaviruses (e.g., Influenza A). There have also been recurring regional outbreaks of mosquito borne diseases caused by RNA viruses such as alphaviruses (e.g., Chikungunya virus) and flaviviruses (e.g., Zika virus, Dengue virus and Yellow Fever virus). These viruses pose a serious threat to human health – in the United States alone, hundreds of thousands of people are hospitalized each year from complications of respiratory viral infections (Thompson et al. 2004). Respiratory viral illnesses are also associated with a steep economic burden; they are collectively estimated to cost over $127 billion each year (Fendrick et al. 2003; Young-Xu et al. 2017).

Unbiased, metagenomic sequence-based approaches could lead to early detection of both previously characterized and novel RNA viruses and serve as a universal assay to detect infectious disease agents (Bibby 2013; Miller et al. 2013; Schlaberg et al. 2017). Established clinical tests largely rely on PCR, culturing or antibody specificity to detect pathogens. These approaches are limited in that they require prior assumptions about the pathogens that might be present. Furthermore, many diagnostic tests rely on culturing the pathogen, which can create delays of several days to diagnosis. Hypothesis-free approaches have the distinct advantage of being able to survey the presence of multiple infectious agents at once in a single assay, which allows pathogens to be identified more quickly and without collecting further samples for a series of tests.

Nanopore sequencing is a cost-effective third-generation sequencing method that allows long reads to be generated on a hand-held device. It has a shorter turn-around time than other next-generation sequencing technologies (Petersen et al. 2019; Miller and Chiu 2022) and has been successfully employed to identify viral (Russell et al. 2018; Arévalo et al. 2022), fungal (Ohta et al. 2023) and bacterial agents (Hewitt et al. 2017; Charalampous et al. 2019; Bouchiat et al. 2022). Nanopore sequencing, unlike traditional RT-PCR methods to diagnose viral illness, allows for whole genome sequencing of viruses, allowing the identification of viral variants to improve disease surveillance efforts. This was used in the recent COVID-19 pandemic to monitor emerging SARS-CoV-2 variants (Yakovleva et al. 2022; Centers for Disease Control and Prevention 2022). To effectively bring this sequencing to a clinical diagnostic setting, there is a need to effectively validate the use of sequencing assays on clinical samples and develop bioinformatics pipelines that can generate reports that provide clinically actionable insights (Miller et al. 2019).

Here, we validate the use of a metagenomic sequencing workflow that rapidly and accurately detects RNA viruses in multiple clinical sample types. This workflow uses unbiased amplification to increase assay sensitivity and multiplexed sequencing to increase throughput and decrease sample analysis cost. Several performance metrics were collected to validate the use of this assay for clinical metagenomics. The limit of detection (LoD) and precision of this assay were determined for seven RNA viruses using contrived samples. We also evaluated the degree to which common contaminants in clinical samples, such as EDTA, interfered with pathogen detection in this workflow. Furthermore, this workflow was tested on commercially purchased remnant clinical samples in addition to contrived samples created in our laboratory, allowing evaluation of assay performance directly against existing clinical assays to determine accuracy.

## Results

### Sample Processing and Bioinformatics Analysis

We developed an untargeted third-generation sequencing assay for RNA virus pathogen identification from nasopharyngeal swabs in viral transport media (VTM) and plasma (Figure 1). This assay includes library preparation, sequencing, and bioinformatics analysis. We then evaluated the performance of the assay using contrived and clinically relevant remnant samples. For each sequencing run, internal sequencing controls consisting of MS2 Phage were spiked into each of the six clinically relevant samples and a NTC (“no template control”) of sterile phosphate buffered saline processed in parallel through RNA extraction. Each sequencing run also had a PC (positive control) consisting of an RNA transcript of the *gfp* gene which was processed through the RNA sequencing library preparation in parallel with the RNA extracted clinically relevant samples and NTC. The sequencing library preparation (Figure 1) for RNA for the untargeted sequencing assay included (1) double-stranded complimentary DNA synthesis (dscDNA synthesis), (2) whole-genome amplification (WGA), (3) ONT native barcoding kit, (4) library pooling in equal molar concentration, (5) ONT ligation sequencing kit, and (6) sequencing on the MinION^TM^ (Mk1B or Mk1C). Raw reads, ‘squiggles’ in the fast5 format, were basecalled and demultiplexed either ‘live’ while sequencing or post-run basecalled using ONT’s official ‘guppy’ basecaller. The resulting demultiplexed and quality filtered reads were analyzed through a bioinformatic pipeline using Epi2Me-Labs as a framework. The SigSciDx bioinformatic pipeline consisted of mapping each of the barcoded read bins to the human genomic sequence using ‘minimap2’ (Li 2021, 2018) to remove human aligned reads. The remaining reads were then mapped to the positive control gene sequence (*gfp*) and internal sequencing control (MS2 Phage) using ‘minimap2’ and alignment statistics were generated with ‘samtools’ (Danecek et al. 2021; Li et al. 2009) for assessment of quality of the sequencing run and for the individual samples. The barcoded read bins were also analyzed for taxonomical determination through the ‘reference inference’ module from SeqScreen. Results from the SigSciDx bioinformatic pipeline were published in a html formatted report for clinician review and interpretation. All remnant samples, positive or negative, were blinded to the analysts performing the extraction and library preparation, and to the analysts performing initial bioinformatic evaluation and interpretation.

**Figure 1.**
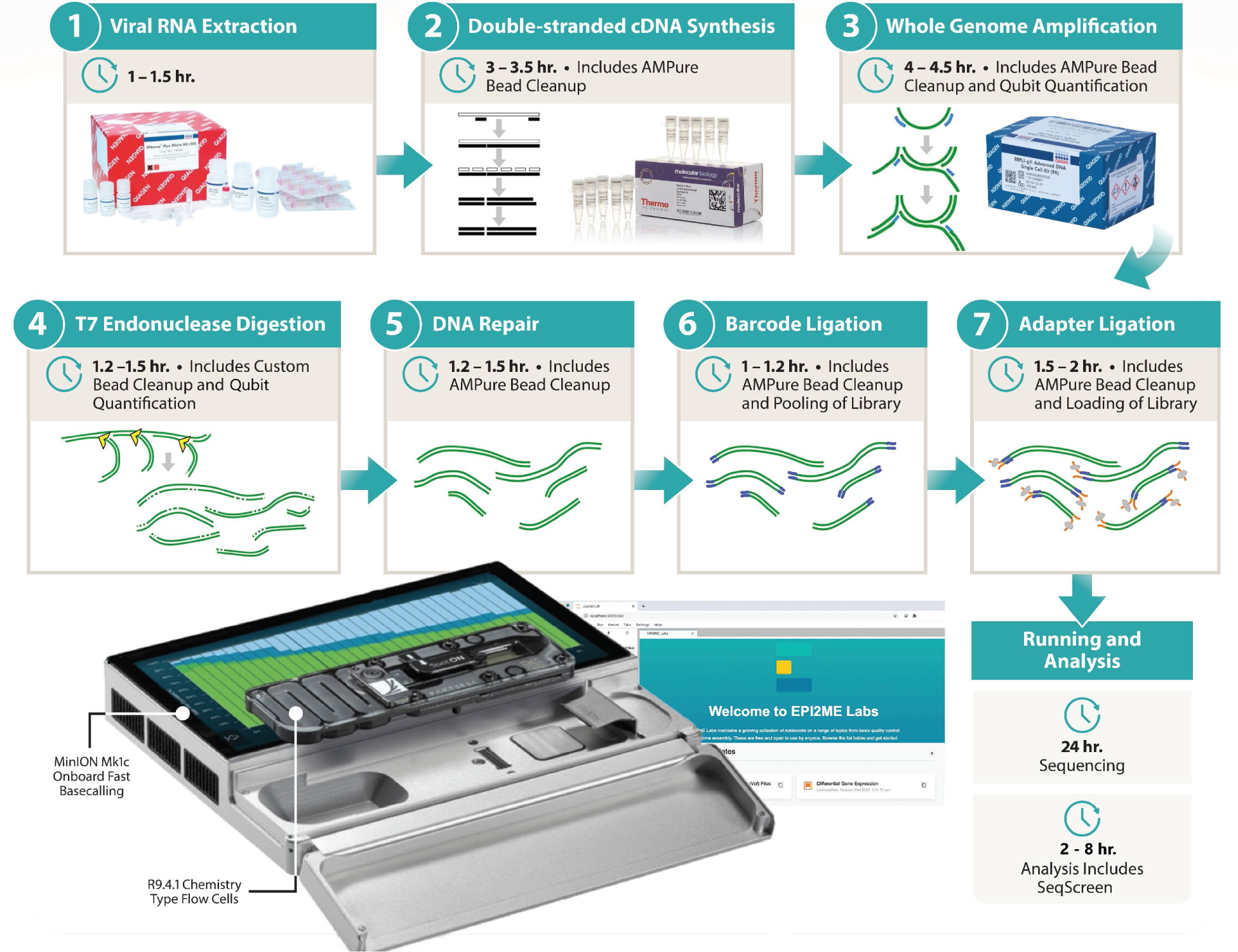
Untargeted sequencing workflow.

### Establishing thresholds for reporting detected pathogens

To minimize false-positive results from cross-contamination caused by low-level barcode crosstalk (Xu et al. 2018), we examined the use of ONT’s current official basecaller ‘guppy’ on 1) a setting for single-end barcode binning of reads, requiring detection of only one barcode per sequence read, or 2) both-end barcode binning, requiring matching barcodes are present at both ends of each read. We examined crosstalk from samples in the NTC (Figure 2, Table S1), indicating potential false positives and the loss of sensitivity (Table 1).

**Figure 2.**
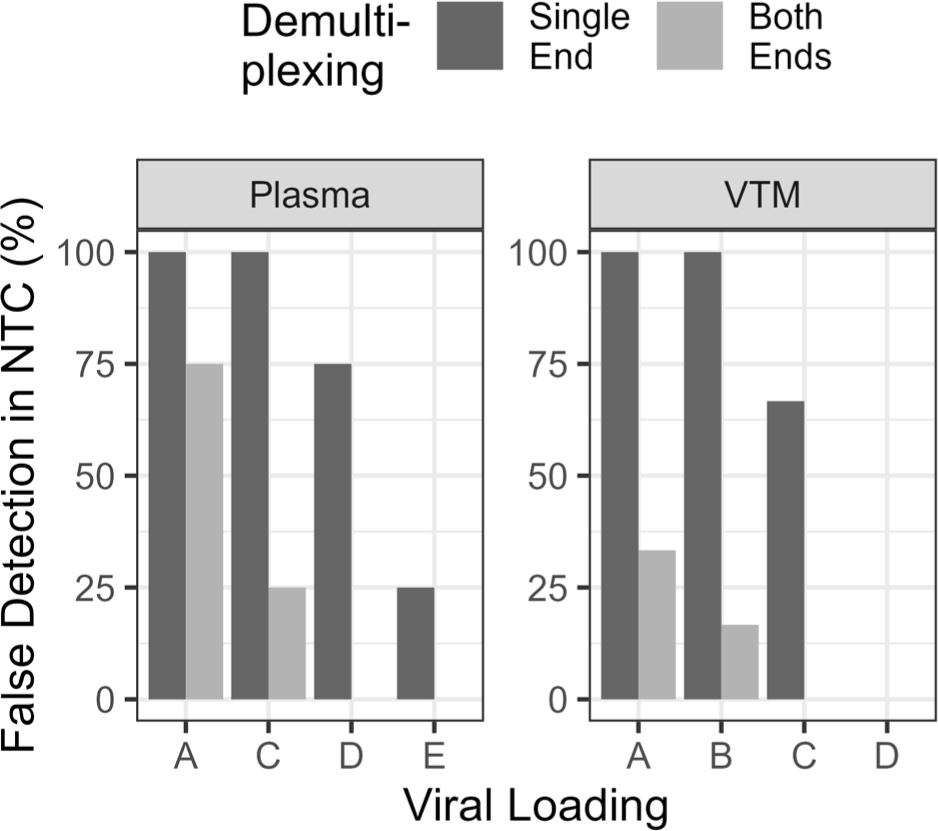
Indication of Barcode Crosstalk by Detection of RNA Viruses in NTC of a Run. Percent of RNA viruses detected in contrived samples at different loadings (**Table 4**) across run using single-end or both-ends demultiplexing methods.

**Table 1.**
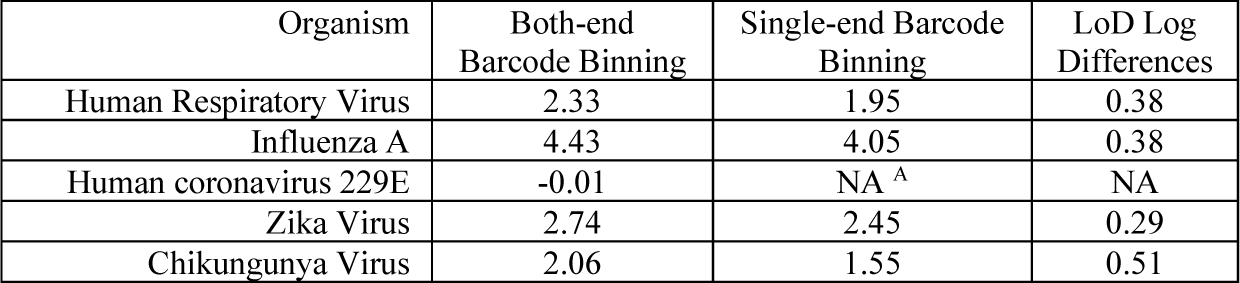
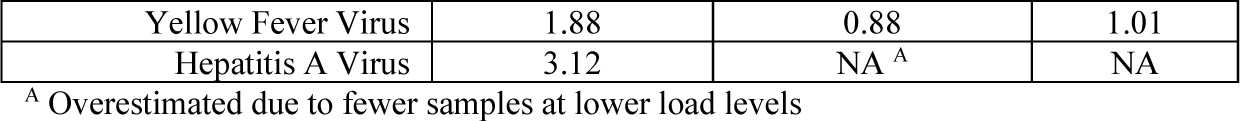
Comparison of Limit of Detection (LoD) Differences between Single and Both-end Barcoding Binning.

Utilizing the both-end barcode binning setting on the basecaller significantly reduced inherent potential false positive calls within the NTC with only a minor reduction in sensitivity of between 0.3 and 1-Log. Total read counts of binned barcodes reduced crosstalk by 38.9 ± 5.5 % between single-end and both-end binning settings.

The sequence analysis results from contrived and remnant clinical samples informed the SigSciDx bioinformatic workflow thresholds as described in the Materials and Methods. This included limiting the number of taxonomical ids initially entering the ‘reference inference module’ and decreasing the stringency of which reference genomic sequences proceed from the first round of mapping to the second round of mapping within the ‘reference inference module’. The criteria for analyst interpretation of detection results were based on replicate sequencing results and inter-run information from contrived and remnant clinical samples, including those from PC, NTC, and internal control. Total number of reads per a barcode with a quality score greater than Q8 was expected to be greater than 40,000. The MinION^TM^ run was expected to achieve greater than 1.5 million long-reads representing greater than 4 Gb estimated bases with an estimated N50 greater than 2.5 kb, usually 3.5 kb. The minimum number of *gfp* gene reads in the PC representative barcode must have been greater than 5,000 for a successful run to be considered for further interpretation. MS2 Phage was accessed on two separate criteria based on mapped read count or percentage, depending on which was greater. MS2 Phage in each sample was expected to have mapped greater than 1000 reads, especially in samples with many reads remaining after host removal, or at 50% of the reads remaining after host removal, especially if fewer reads remain after host removal. The NTC and *gfp* gene barcode samples were used to interpret if high barcode crosstalk or cross-contamination exist. It was expected that only MS2 Phage, or other phages, may be detected within the NTC. If an NTC detected a virus that was also detected in at least one other sample of the run, this dictated a deeper review. Generally, the preferred remedy was determining the sample with a high read count which was positive for the detected virus and re-sequence using the cleaned WGA DNA material of the other samples to verify their positive calls. Otherwise, resequencing all the samples again was also viable. There was little ability to access if a statistical cut-off could be employed such as requiring a 2 or 3 -fold higher number of reads in a sample compared to the NTC for that virus to indicate a detection, as this appeared in only 1 of the runs performed on the remnant samples. Further study of remnant or clinical samples could inform a statistical required read count in the presence of barcode crosstalk or cross-contamination.

### Limit of Detection

A 95% limit of detection (LoD) was determined for each of the three and four representative RNA viruses in VTM and plasma contrived samples, respectively. We evaluated each RNA virus over a minimum 6-Log dilution range, testing 2 to 14 replicates at each concentration. The 95% LoD, defined as the lowest concentration at which 95% of positive samples are detected, was determined for each of the seven RNA viruses using probit analysis (Table 2).

**Table 2.** Performance characteristics for the untargeted sequencing assay.

| Performance metric | Method | Result |  |  |
| --- | --- | --- | --- | --- |
| Limit of detection (LoD) | Qualitative detection of RNA virus dilution replicates by probit analysis | Pathogen | LoD (Estimated PFU<sup>1</sup>) |  |
|  |  | Influenza A Virus | 26,915 TCID<sub>50</sub>/mL (18,840 PFU/mL) |  |
|  |  | Human Respiratory Syncytial Virus | 214 TCID<sub>50</sub>/mL (150 PFU/mL) |  |
|  |  | Human Coronavirus 229E | 1 TCID<sub>50</sub>/mL (1 PFU/ mL) |  |
|  |  | Zika Virus | 550 TCID<sub>50</sub>/mL (385 PFU/ mL) |  |
|  |  | Hepatitis A Virus | 1318 TCID<sub>50</sub>/mL (923 PFU/ mL) |  |
|  |  | Yellow Fever Virus | 68 TCID<sub>50</sub>/mL (47 PFU/ mL) |  |
|  |  | Chikungunya Virus | 115 TCID<sub>50</sub>/mL (80 PFU/ mL) |  |
| Precision | Qualitative detection over 2 to 7 contrived sample runs of each organism (inter-assay)<sup>2</sup> | 100 % concordance |  |  |
|  | Qualitative detection of duplicate contrived samples on the same run (intra-assay)<sup>2</sup> | 100 % concordance |  |  |
|  | Qualitative detection over 21 remnant sample (duplicate or triplicate) runs (inter-assay) | 100% concordance |  |  |
| Interference | Quantitative read count of viruses with spiked blood (5%, 2%) | Decrease of number of reads by 1.3-Logs to 1.5-Logs mapped to viruses |  |  |
|  | Quantitative read count of viruses with spiked EDTA (100 mM, 10 mM, 5 mM) | Decrease of number of reads by 0.6-Logs and 0.1-Logs with addition of 100 mM and 10 mM EDTA, respectively |  |  |
|  | Quantitative read count of viruses with spiked bacteria (*Micrococcus luteus* or *Staphylococcus epidermidis*) | No significant changes in number of reads mapped to RNA viruses |  |  |
| Accuracy | 18 of single-detection remnant sample runs, results comparisons (3 for each organism). |  | <u>Sensitivity</u> | <u>Specificity</u> |
|  |  | Human metapneumovirus | 33.3%<sup>3</sup> | 100% |
|  |  | Parainfluenza IV | 33.3% | 100% |
|  |  | SARS-CoV-2 | 100% | 100% |
|  |  | RSV | 100% | 100% |
|  |  | Influenza A | 100% | 100% |
|  |  | Enterovirus | 100% | 100% |

<sup>1</sup> Estimated PFU calculated by multiplying the TCID<sub>50</sub> /mL by 0.7<sup>2</sup> Nearest higher loading (Table 4) to that of the LOD (within 1-Log)<sup>3</sup> Includes two of the three samples that did not pass internal control QC

### Precision

We demonstrated inter-assay reproducibility of the untargeted sequencing assay by testing of the NTC and contrived samples at the nearest loading above the calculated LoD for each organism across at least 4 sequencing runs and intra-assay reproducibility by testing of at least 4 independently generated sets of duplicate contrived samples on the same run (and negative remnant samples in two runs). The *gfp* gene mRNA transcript positive control passed QC for every completed sequencing run (36 total runs: 20 contrived sample runs and 16 remnant sample runs), indicating successful sequencing library generation from cDNA synthesis through sequencing. Internal spiked MS2 phage controls passed QC for all sequenced replicate contrived samples (216 total), indicating conditions across all processes of the workflow were successful, including the RNA extraction process. For remnant samples, 2 samples consistently failed internal spiked phage control QC after 3 independent runs were performed (6 total remnant replicate samples). The remaining 90 of the 96 replicate remnant samples across the 16 runs passed QC for the internal spiked MS2 phage control. The 6 failed internal controls in the replicates of the 2 remnant samples occurred in samples that were Human metapneumovirus-positive by original clinical assay and had relative low human sequence contributions compared to the other VTM samples. One of the samples, despite the failed QC, detected the presence of Human metapneumovirus while the other consistently failed to detect. All seven organisms in the contrived samples were detected using pre-established threshold criteria for the duplicate intra-assay samples on the same run and each replicate inter-assay run (Table 2). All 9 negative remnant samples, determined to be absent of viral pathogens by previous clinical testing, had no detection by untargeted sequencing of pathogenic RNA viruses within the same run which were divided between 2 individual runs (intra-assay testing). From across all 36 sequencing runs (inter-assay testing), 35 NTCs showed no pathogenic RNA virus detections. The single run that showed a positive detection within the NTC, presence of Influenza A, was a remnant run containing a patient sample with a high-load of Influenza A and subsequent two repeats of the run using that sample did not show detection in the NTCs. The read count was at 14 for Influenza A within the NTC and had enough coverage and depth to be determined as present despite a lower calculated statistics for the mapping. Interpretation was not impacted with the other remnant samples in the run based on pre-established interpretation guidelines and criteria.

### Accuracy

A total of 31 remnant samples consisting of 18 samples with original clinical positive single detections and 13 samples with negative clinical detections were tested using the untargeted sequencing assay. Concordance was determined by comparing the assay results to original clinical test results (Table 2 and Table S2). Two clinical positive samples failed internal control QC in 3 independent runs. Of the other 16 clinical positive samples, 13 showed positive concordant detections or true-positives and 3 samples showed unexpected negative detections and were classified as false-negatives for detection by the untargeted assay. All 13 negative clinical samples were also negative for the untargeted sequencing assay.

Overall, the untargeted sequencing assay showed 81.25 % sensitivity (13/16) and 100% specificity (29/29) compared to original clinical positive sample results excluding the internal control QC failures (Table 2). No cases, excluding the internal control QC failed samples, were classified as untargeted sequencing assay false positives and two samples originally identified as Parainfluenza IV were classified as false negatives; discrepancy testing was not performed.

### Interference

We evaluated the effects of interference from blood, EDTA, and increasing additions of *Micrococcus luteus* or *Staphylococcus epidermidis* to contrived plasma samples prior to extraction on the untargeted sequencing assay. Exogenous addition of blood to plasma samples at 2% and 5% of the sample prior to RNA extraction resulted in a significant 1.34 ± 0.04 -Log and 1.54 ± 0.11 -Log reduction in the number of RNA virus reads, respectively (Figure 3) ANCOVA and TukeyHSD, p<0.001). The MS2 Phage internal control for the samples indicated only a 0.83-Log reduction in reads (Figure 3).

**Figure 3.**
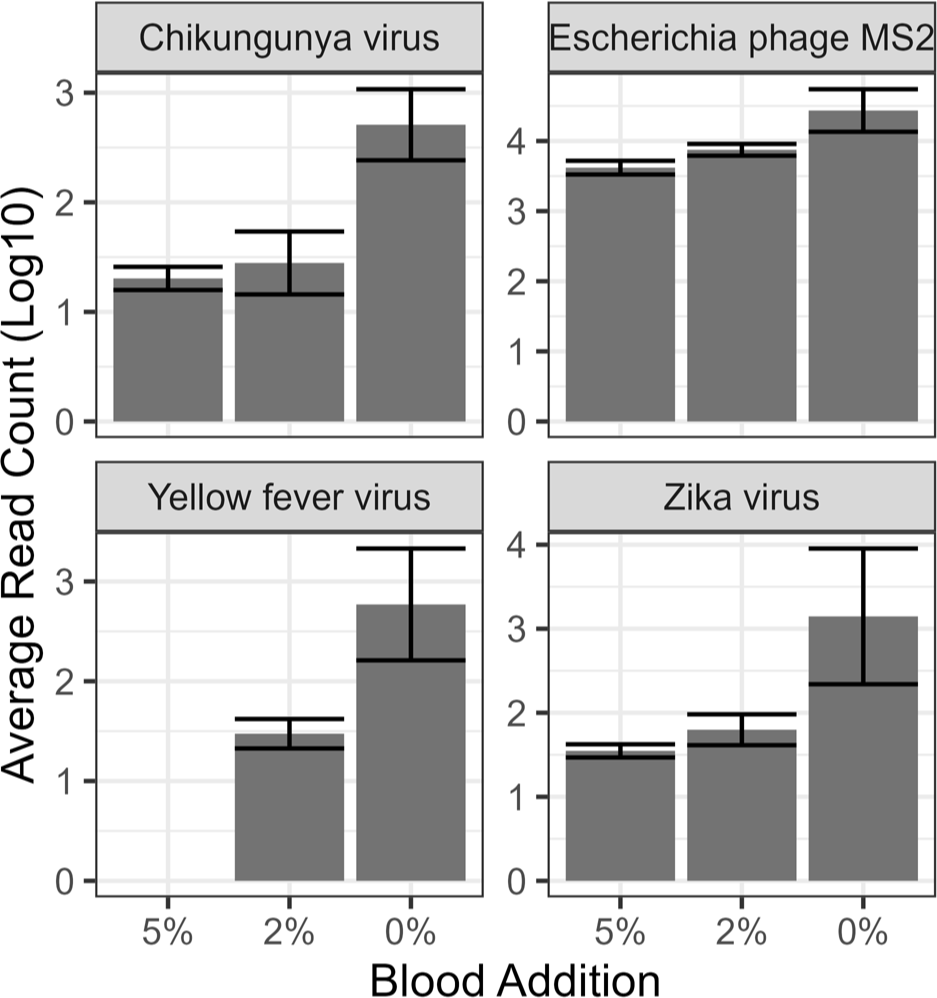
Interference of RNA virus reads with increasing blood concentration. Number of reads mapped to the individual viruses within contrived samples containing increasing additions of human whole blood.

The Log reduction in the number of RNA virus reads was also consistent with increasing concentrations of virus within the sample containing 2% blood (Figure 4). The addition of 2% blood to samples with increasing concentrations of viral pathogens had a significant reduction of 1.31 ± 0.05 -Log reads at all pathogen concentrations, with the exception of ‘low’ pathogen concentrations where the read count dropped below the 10 read threshold and was removed from analysis (ANCOVA and TukeyHSD, p<0.001). The MS2 Phage internal control had only a 0.5-Log reduction in reads with greater variability in the ‘high’ load viral pathogens in 2% blood.

**Figure 4.**
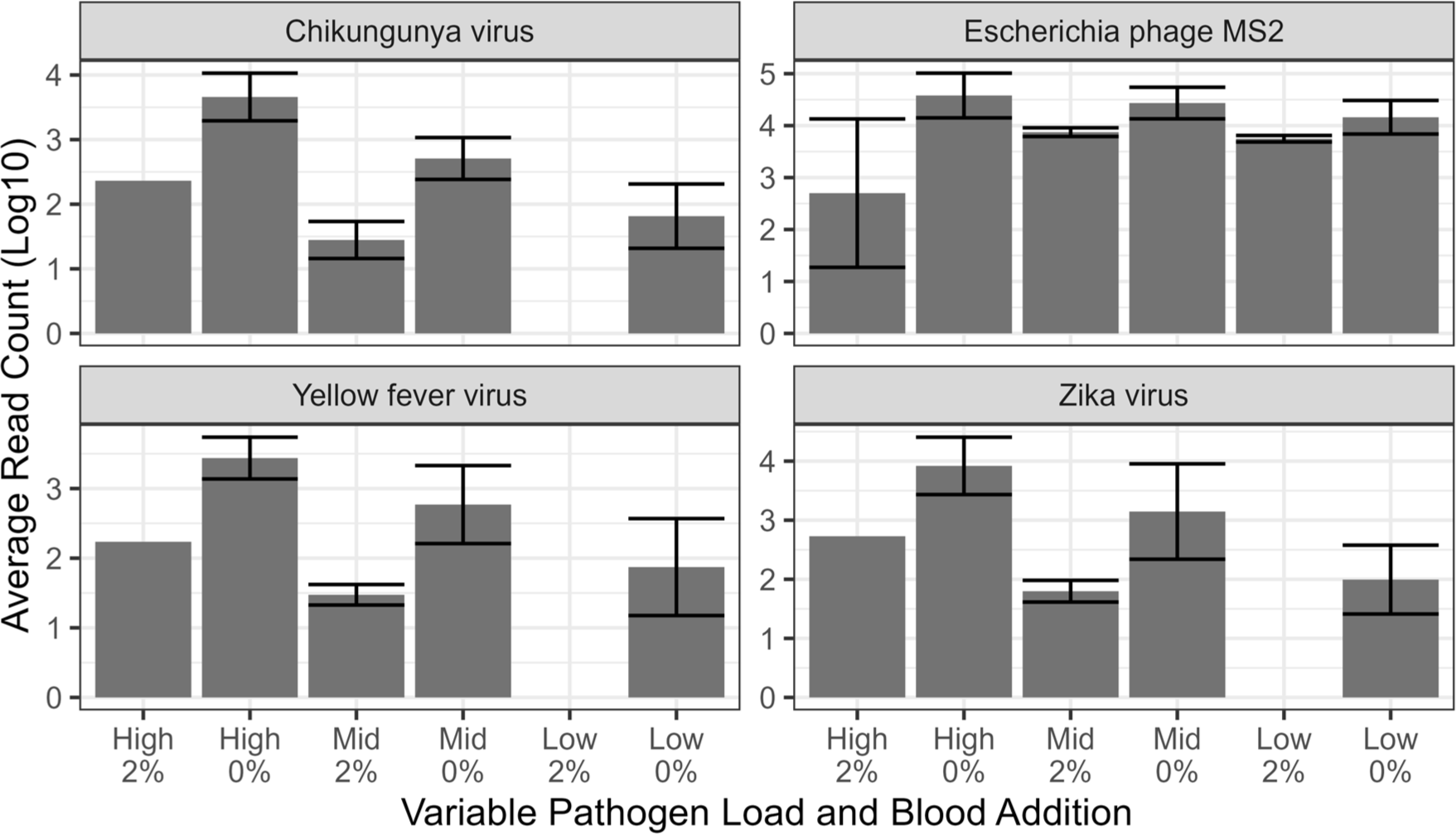
Interference from blood with increasing RNA virus concentration. Number of reads associated with individual viruses at different loading in contrived samples in the presence of 2% blood. Threshold for detection was a minimum of 10 reads leading to absence in graph and subsequent statistical analysis.

Interference caused by the addition of increasing concentration of EDTA was also established (Figure 5). The addition of low concentrations of EDTA as sodium salt at 5 mM to the existing approximate 4.5 mM potassium EDTA for plasma collection may have increased the extraction efficiency of some RNA viruses with an approximate increase in viral reads by 0.3-Log, yet was not statistically significant compared to no EDTA addition (ANCOVA and TukeyHSD, p=0.69). This potential increase was eliminated at addition of 10 mM EDTA levels (p=0.99 compared to no addition) and negatively impacted RNA virus reads by approximately 0.60 ± 0.13 -Log reduction at additions of 100 mM EDTA compared to no addition (p=0.005).

**Figure 5.**
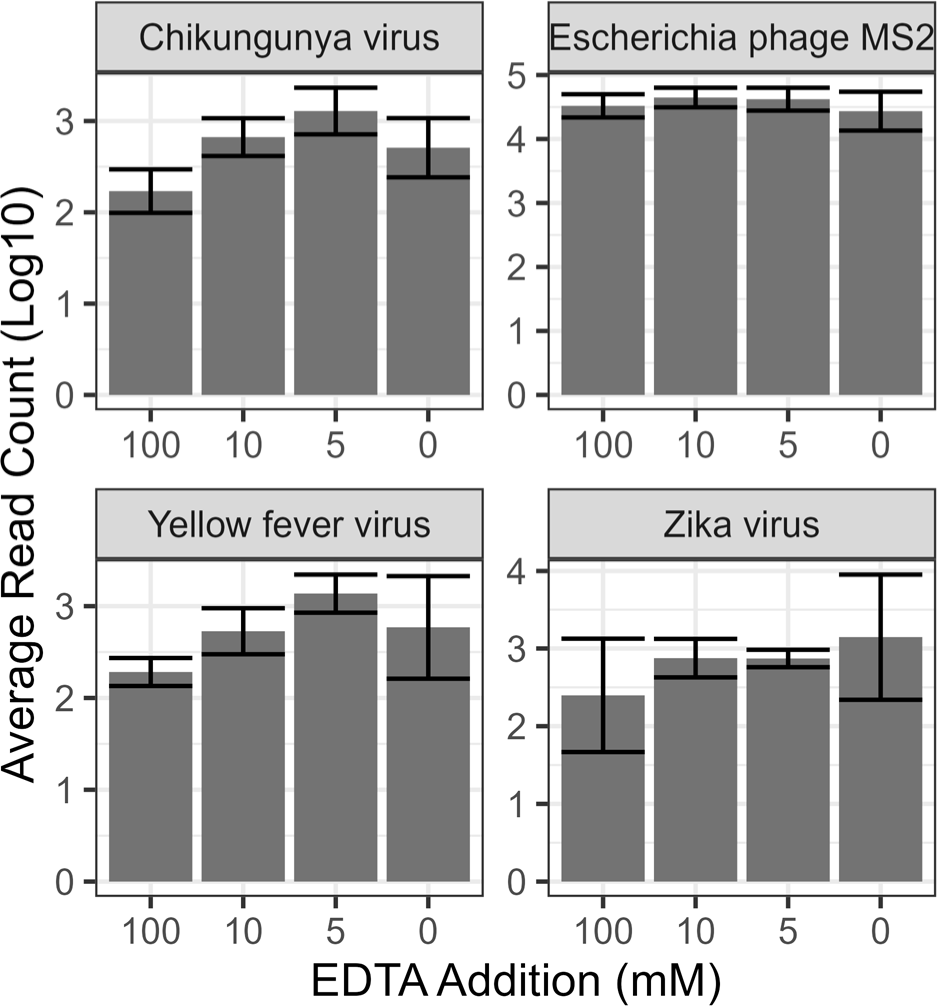
Interference in the presence of EDTA. Number of reads associated with different viruses within contrived samples with increasing concentrations of EDTA.

Finally, we evaluated the addition of *Staphylococcus epidermidis* or *Micrococcus luteus* to known positive plasma samples, neither of which negatively impacted viral read counts (Figure 6, ANCOVA: p=0.239).

**Figure 6.**
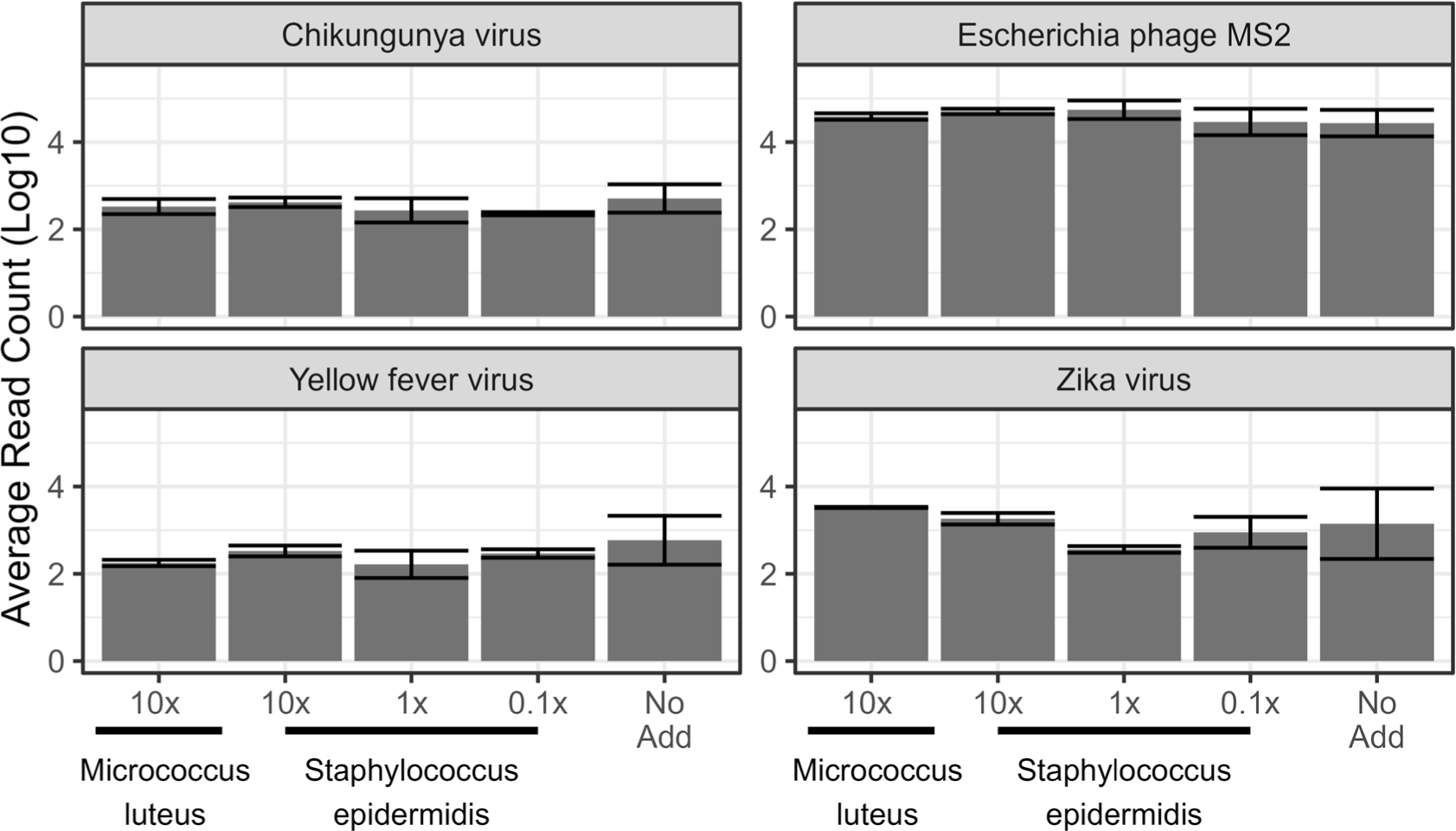
Interference by bacteria addition. Number of reads associated with different viruses in contrived samples with increasing concentrations of *Staphylococcus epidermidis* or *Micrococcus luteus*

The relative selectivity of sequences within the different clinical sample matrices, VTM and Plasma, and the absence of a complex matrix, NTC, was examined using the MS2 Phage IC used between the samples ( Figure 7). There was no significant difference in the number of MS2 Phage reads between the remnant and contrived samples runs (ANCOVA, p=0.537). There was also no significant difference in the number of MS2 Phage reads between the plasma and VTM in either the remnant (TukeyHSD, p=0.18) or contrived (TukeyHSD, p=0.78) runs. The number of MS2 Phage reads in VTM samples had a significant decrease of 1.13 and 1.16-Log compared to NTC (TukeyHSD, p<0.001) in contrived and remnant samples respectively. Plasma had a significant decrease in MS2 Phage reads of 0.92-Log in contrived sample runs (TukeHSD, p<0.001) but not in remnant sample runs (0.68-Log; TukeyHSD p=0.086) compared to NTC. There was no significant difference between the number of human reads between the contrived and remnant samples (ANCOVA, p=0.66) nor between VTM and plasma (ANCOVA, p=0.99). The presence of human reads in the NTC and PC of approximately 300 reads indicates how the majority reads from 6 samples can impact the contamination of reads in samples expected not to contain human reads.

**Figure 7.**
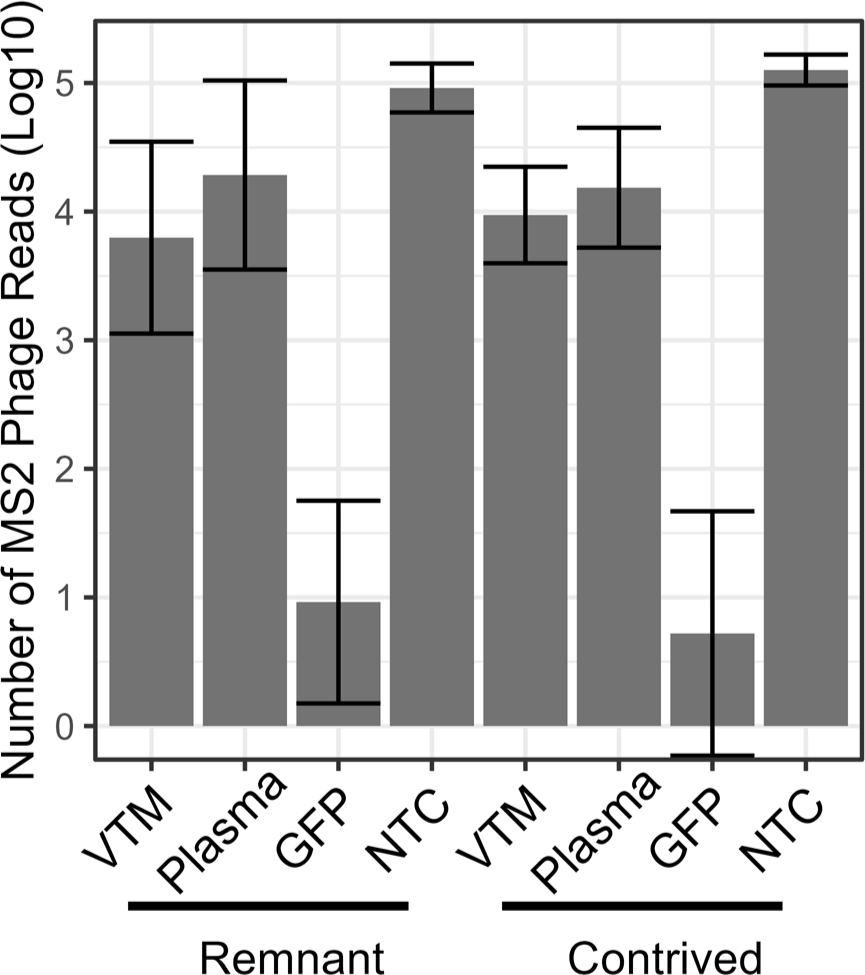
Number of MS2 Phage Reads in Different Sample Matrices.

### Challenge remnant samples

We evaluated remnant samples containing multiple virus detections by current molecular methods or non-overlapping single calls (Table 3). Of the 17 diagnosed viruses present by current molecular diagnostics in the 9 samples, 8 of those viruses were confirmed through untargeted sequencing (47%) without incorrect identification of other viruses. Discrepancy testing was not performed.

**Table 3.**
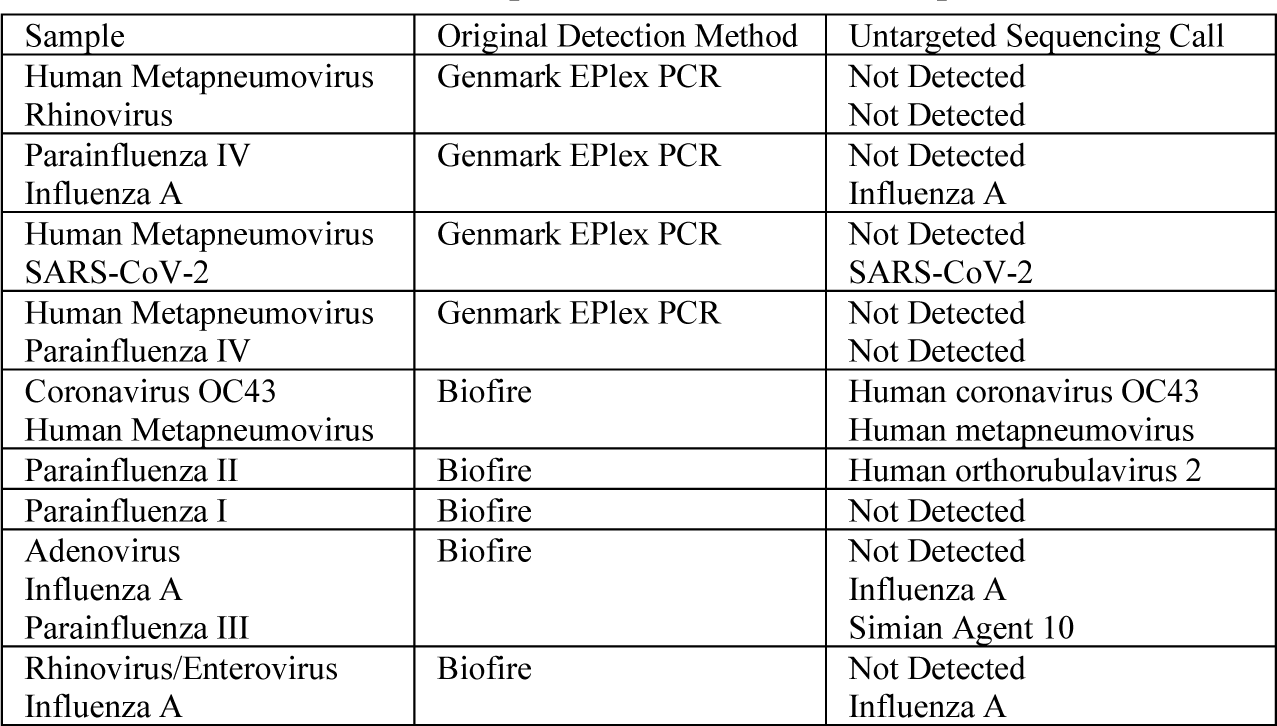
Multiple-detection remnant samples.

## Discussion

We have previously developed and evaluated a clinical untargeted sequencing assay intended to aid in the diagnosis of respiratory and blood-borne infections by RNA viruses (Kappell et al. 2023). Here, we rigorously evaluate the best performing of the sequencing methods for different RNA viruses within viral transport media and human plasma through clinically relevant contrived samples and remnant patient samples. We incorporated multiple QC materials, including an external positive control (*gfp* gene transcript RNA), NTC samples that are run in parallel with clinical samples, and spiked MS2 Phage as an internal process control. Reproducible threshold metrics were established and evaluated to enable identification of pathogens from long-read nanopore sequencing data above background noise and to minimize potential barcode crosstalk within multiplexed analyses. The final sequencing protocol incorporated: (1) RNA extraction of MS2 Phage spiked samples and NTC, (2) reverse transcription of RNA into double-stranded cDNA, (3) unbiased amplification through whole genome amplification, (4) ligation of barcodes and adapters using ONT kits, (5) sequencing, basecalling, and demultiplexing using ONT MinION™ (Mk1B or Mk1C), and (6) SigSciDx open-source bioinformatics pipeline for QC and pathogen detection.

Our untargeted sequencing library preparation protocol uses a whole genome amplification (WGA) approach after cDNA synthesis. This step increases assay specificity without requiring targeted amplification. The WGA step has the potential to be further optimized to shorten the time of reaction, but the material generated was sufficient for approximately 3 reactions after a 2-hour incubation, starting with nearly undetectable quantities of cDNA. The amplified material serves as an ideal reference point for additional sequencing or molecular analysis, if needed. WGA was performed using Qiagen REPLI-g kit which has been shown to introduce the lowest amplification bias compared to other multiple displacement amplification (MDA) methods (Pinard et al. 2006) inducing the least distortion in read counts per bin across the length of a genome compared to unamplified controls. While sequencing independent single primer amplification (SISPA) methods are generally quicker than MDA methods, MDA methods are known to perform better than SISPA methods in sequencing output, taxonomic assignments, diversity, and assembly statistics (Kallies et al. 2019; Parras-Moltó et al. 2018). The simplified execution of a MDA based method in the Qiagen kit format has led to our adoption of the method for this untargeted sequencing assay.

Contrived VTM and plasma samples in this study that contained 3 and 4 RNA viruses respectively, indicated a range of limits of detection for the untargeted sequencing assay ranging from 1 TCID_50_ /mL (est. 1 PFU/ mL) for Human coronavirus 229E to 26,915 TCID_50_ /mL (est. 18,840 PFU / mL) for Influenza A virus. Untargeted sequencing assay sensitivity for detection of a given organism is dependent on multiple factors, including extraction efficiency, size of the genome, complexity of the genome, the variability of genomic equivalences (sequenceable material) vs infectious particles, library preparation bias, and availability of matching reference genomes in the database. We think the higher levels of detection for Influenza A are likely due to genome complexity and size. While Influenza A has an approximately 13.5 kb genome, it is fragmented into 8 segments ranging in size from 890 to 2,341 bp. The subset of Influenza A genomes available in the NCBI Genbank of complete genomes is also limited, preventing accurate strain level detection. We speculate that Human coronavirus 229E was detected at lower LOD levels due to its large unsegmented 27.5 kb genome allowing multiple targets and ease of progressivity for reverse-transcription and WGA. Another important context of the LOD for the individual viruses used in the study is the variability of genome equivalents infectious particles in a viral stock, similar to particle-to-PFU ratios (Bhat et al. 2022). For example, influenza has a particle-to-PFU ratio of 20-to-50 and SARS-CoV-2 has a range of 10^4^-to-10^6^ for genomic RNA to PFU. As viral stocks used for this study were reported in TCID_50_ related to PFU, correcting for the particle-to-PFU ratio consistent with these values would suggest a more consistent LOD between these viruses.

A key limitation for infectious disease diagnostics using untargeted sequencing assays is background interference. The presence of blood as little as 2% volume of the sample led to a greater than 10-fold decrease in the number of reads, indicating a potential 1-Log decrease in analytical sensitivity of the assay. This highlights the importance of the plasma capture step from a blood sample during initial sample preparation to maximize assay sensitivity. Additionally, the presence of 100 mM EDTA caused a 0.6-Log reduction in reads and potential loss in analytical sensitivity. The presence of such an excess of EDTA is unlikely, but a low volume of blood captured in a K_2_-EDTA vacutainer tube would increase the concentration of EDTA present in the extracted sample could be indicated as a potential cause for sequencing failure. The spiked MS2 phage IC was useful for assessing the decreased assay sensitivity caused by interferents or due to matrix effects. Notably, matrix effects on read counts from the MS2 Phage IC were more variable in the remnant samples compared to the NTC. While these matrix effects had an impact on total read output, the read count following host removal was most noteworthy. Remnant samples that had fewer mapped MS2 reads following host removal typically showed fewer or no pathogen reads. This indicated potential interference from high human background or potentially other confounding inhibitors. Accordingly, repeated failures or low read abundance of the MS2 Phage IC may indicate poor assay performance. In this case, other diagnostic tests that are less sensitive to interference should be considered (e.g., RT-PCR).

Of the 35 RNA viruses detected by conventional clinical molecular assays across 27 total remnant samples, untargeted sequencing detected 22 concordant positives. Concordant negative detections occurred in 13 out of 13 negative remnant samples. These findings show the unbiased sequencing assay has 62.9% sensitivity and 100% specificity. The calculated 62.9% sensitivity refers to clinical sensitivity in diagnosis of infection and not analytical sensitivity or detection of pathogen nucleic acid. Multiple factors likely play a role in limiting the clinical sensitivity of untargeted sequencing, including: (1) the use of remnant samples as a “gold standard”, with the potential that some samples may represent false positives due to PCR artifacts or incorrect handling (e.g., contamination), (2) the potential that remnant samples have undergone nucleic acid degradation from prior freeze-thaw steps, (3) the necessary use of both-end barcode demultiplexing and robust pre-established thresholds to minimize false-positive detections that could filter appropriate pathogen sequencing data, and (5) potential patient medications, treatments, or other interferents that cause assay inhibition. Discrepancy testing was not performed to determine the potential causes for the false-negative cases.

Approximately 14 of the 40 total remnant samples used in this study, including negative remnant samples, had additional virus species or genera detected above pre-established thresholds consisting of environmental or normal flora. Most of the additional detections were of phage, most likely due to the mappings of the MS2 Phage to additional phage sequences and small amounts of contamination in workflow reagents. Torque teno viruses, Torque teno midi virus, and TTV-like mini viruses were also identified in several of the remnant VTM samples. These anelloviruses have small circular, negative-sense, single-stranded DNA genomes which may have escaped the DNA removal column-based step in the RNA extraction process. These anelloviruses are not directly concerning as they are diverse and commensal members of the human virome (Bendinelli and Maggi 2010). However, their increase leading to detection may indicate host immunosuppression (del Rosal et al. 2023; Prasetyo et al. 2015). Determining the clinical significance of detecting viruses that may be contaminants or normal flora remains a classical problem in clinical microbiology and often requires clinical context for interpretation. While plasma is normally a sterile sample, manipulation and handling of the sample may introduce low levels of contamination which ultimately gets amplified by our use of WGA within the untargeted sequencing protocol. Additionally, the collection using nasopharyngeal swabs into VTM in a non-sterile site collection environment presents a reasonable risk of several potential environmental and normal flora being captured along with viral pathogens. Thus, detection of multiple viruses as potential contamination or normal flora were noted and were considered negative in our evaluation of assay performance.

Untargeted sequencing testing can provide broad-spectrum RNA virus detection and identification, however assessment of the clinical significance of the reported findings may require interpretation. Clinical context of the results from untargeted sequencing is important in discussing and reviewing patient cases with the treating clinicians. The potential for untargeted sequencing for RNA virus identification includes characterization of antiviral or vaccination escape mutations, genotyping or strain-level identification, and presence of reads from potential pathogens below formal threshold that can inform follow-up targeted testing.

## Methods

### Remnant Clinical Samples

Clinically diagnosed, de-identified remnant samples were obtained from Discovery Life Sciences and Precision for Medicine. Of the 40 remnant samples evaluated, 9 VTM samples were negative based on molecular respiratory panels, 4 were negative serum or plasma samples, and 27 VTM samples were positive for single or multiple respiratory viruses. Of the remnant samples, 6 different RNA virus pathogens as determined by standard clinical testing were represented by 3 samples each. These 18 remnant VTM samples were used for the initial accuracy study. The other 9 VTM samples containing mixed or singlet samples with one diagnosed pathogen were kept for additional challenge samples. Remnant samples were pipetted into 140 µL single use aliquots for extraction. The number of prior freeze thaws of remnant samples was unknown before receipt. Samples were stored at -80 °C upon receipt and again after aliquots were prepared.

### Contrived Sample Preparation

Aliquots of negative diagnosed remnant VTM or plasma were pooled to generate a matrix for contrived RNA virus samples. Contrived samples in VTM were prepared using Influenza A virus (BEI Resources NR-42007), Respiratory Syncytial virus (BEI Resources NR-28529), and Human coronavirus (HCoV) 229E (NR-52726). Contrived samples in plasma were prepared using Chikungunya virus (NR-56523), Hepatitis A virus (BEI Resources NR-137), Yellow Fever virus (BEI Resources NR-116), and Zika virus (BEI Resources NR-50065). To determine LoD, contrived samples were prepared at a starting dilution of 25 µL/mL stock material in pooled VTM or plasma. Tenfold serial dilutions were prepared in respective pre-screened negative matrices to a final concentration of 0.00025 µL/mL. The viral load levels in TCID_50_/ mL at the different serial dilutions are indicated in Table 4. All samples were prepared into single use 140 µL sample aliquots and frozen at -80 °C until extraction.

**Table 4.** Contrived Sample Loading.

| Media: |  | Viral Transport Media |  |  | Human Plasma |  |  |  |
| --- | --- | --- | --- | --- | --- | --- | --- | --- |
|  | Virus: | Influenza A Virus<br>(A/Wisconsin/15/2009 (H3N2)) | Human Respiratory Syncytial Virus<br>(A 1998/3-2) | Human Coronavirus (229E) | Zika Virus (MR 766) | Hepatitis A Virus (HM175/18f) | Yellow Fever Virus (17D) | Chikungunya Virus (181/25) |
|  | BEI Cat# | NR-42007 | NR-28529 | NR-52726 | NR-50065 | NR-137 | NR-116 | NR-56523 |
| Viral Load Levels Log<sub>10</sub> (TCID<sub>50</sub>/mL) | A* | 7.10 | 5.60 | 3.85 | 7.35 | 6.85 | 6.60 | 6.35 |
|  | B | 6.10 | 4.60 | 2.85 | 6.35 | 5.85 | 5.60 | 5.35 |
|  | C | 5.10 | 3.60 | 1.85 | 5.35 | 4.85 | 4.60 | 4.35 |
|  | D | 4.10 | 2.60 | 0.85 | 4.35 | 3.85 | 3.60 | 3.35 |
|  | E | 3.10 | 1.60 | -0.15 | 3.35 | 2.85 | 2.60 | 2.35 |
|  | F | 2.10 | 0.60 | -1.15 | 2.35 | 1.85 | 1.60 | 1.35 |
|  | G | NA | NA | NA | 1.35 | 0.85 | 0.60 | 0.35 |

\*The letters in column 1 provide a designation for the specific viral load of each virus in the corresponding row. Viral loads decrease as the letters continue, with “A” representing the highest load level.

### Untargeted Third-Generation Sequencing

RNA from contrived and remnant clinical samples was extracted using the Qiagen RNeasy Plus Micro Kit (Qiagen #74034) with minor changes to the manufacturer’s standard protocol adapting to the specific sample matrix and input volume of 140 µL sample material augmented with 5 µL of MS2 Phage internal control (ZeptoMetrix, 0810274; 145 µL total input). Extracted RNA was immediately reverse-transcribed using random hexamers and Maxima H Minus Double-Stranded cDNA Synthesis Kit (Thermofisher K2561) following manufacturer’s instructions including the recommended changes to increase the addition of 1st strand enzyme mix to 2 µL and the incubation temperature to 55 °C. In addition to the extracted RNA, an RNA transcript of the *gfp* gene (System Biosciences, LLC MR700A-1) was included as an additional positive control sample at 6 pg. If necessary, DNA forms of samples were stored at -20 °C only after bead clean-up of the previous reaction steps. Double-stranded cDNA was cleaned using AMPure XP beads before whole genome replication using the Qiagen REPLI-g Advanced DNA Single Cell Kit (Qiagen, 150363) following the Qiagen supplementary protocol “Whole genome amplification from genomic DNA using the REPLI-g Advanced DNA Single Cell Kit with increased sample volumes.” Briefly, 15 µL of double-stranded cDNA was denatured with 2 µL of Advanced Buffer DLB for 3 minutes at 25 °C. Stop Solution was added at 3 µL and mixed. The 29 µL REPLI-g sc Advanced Reaction Buffer with 2 µL REPLI-g sc DNA Polymerase was added directly to the denatured cDNA and incubated for 2 hours at 30 °C. The REPLI-g sc DNA polymerase was inactivated at 65 °C for 3 minutes. The resulting amplified DNA was purified from the reaction by AMPure XP bead clean up and digested using T7 Endonuclease I (New England Biolabs (NEB), M0302L). For effective removal of T7 digested fragments, a custom AMPure XP bead solution was made using PEG 8000 50% (w/v) (Fisher Scientific, NC1017553) as described in Oxford Nanopore Technology’s (ONT) whole genome amplification protocol for LSK110 (v110, rev. 10 Nov 2020, Oxford Nanopore Technologies 2023). Once purified, the sequencing library was prepared by using end repair and ligation following the ONT’s protocol for the Ligation sequencing kit (SQK-LSK110). Native barcode expansion kit 1-12 (EXP-NBD104) was used for multiplexing samples. All bead cleanups were done on a microfuge tube magnetic separation stand (Permagen). Sequencing was performed on Oxford Nanopore Technologies MinION^TM^ Mk1B or Mk1C using R9.4.1 flow cells (FLO-MIN106D). Each flow cell was primed and loaded using manufacturer’s instructions. Each run used 24-hour run default settings except no reserve pores. ‘Live’ basecalling was performed in ‘Fast basecalling mode’ and single-end barcode demultiplexing. The ‘live’ basecalling was performed with the latest MinKNOW (22.12.5 to 23.04.03) running the Mk1C or Mk1B which did not impact ‘Fast basecalling’ (guppy 6.4.6 - 6.5.7) based on change logs and no changes on re-basecalling of the earliest run. Alternative basecalling and demultiplexing including both-end barcode demultiplexing of the sequencing runs were performed using standalone ‘guppy’ basecaller version 6.2.1. All data analyzed was basecalled with version 6.2.1 in both-end barcode demultiplexing as a final workflow, with the exception for initial comparison of single-end and double-end barcode demultiplexing (Figure 2, Table 1).

### Bioinformatics Analysis

Analyses were developed using EPI2ME Labs (https://github.com/epi2me-labs), a Jupyter-notebook platform allowing users to run pre-written python, R, or BASH based analyses in a web-browser based environment. For each sequencing run, passing reads (default Q8 threshold) were concatenated for each barcode, and quality of run statistics (e.g., read length and quality distribution) were reported and visualized in the web-based environment. Human reads were removed and reported after mapping to the human genome using ‘minimap2’ (Li 2021, 2018) with default ‘ont’ parameters. The remaining human depleted reads were then mapped to the control sequences of the *gfp* gene and MS2 Phage internal controls using minimap2, while removing the *gfp* mapped reads. The number and percentage of reads aligned to the human genome, *gfp* gene, and MS2 Phage were analyzed using SAMtools (Danecek et al. 2021; Li et al. 2009) and reported in the Quality Control (QC) portion of the output for review. Untargeted sequence analysis was performed using Centrifuge (Kim et al. 2016) with a customized virus database to identify putative NCBI taxonomy identifiers (taxids) and input those taxids into the ‘reference inference module’ of SeqScreen-Nano (Balaji et al. 2023b, 2023a).

The customized Centrifuge database is formed of the ‘Complete Genome’ or ‘Chromosome’ from the viral genomes from NCBI’s GenBank database, excluding all partial or incomplete genome sequences, using the centrifuge-download command. The centrifuge-build command was used as described by Centrifuge developers (Centrifuge Developers 2023; Kim et al. 2016) to generate the custom virus sequence database to use with the Centrifuge taxonomical classifier. The taxids reported by Centrifuge were filtered to include only those taxids with greater than 10 reads assigned to a species or leaf taxonomical level. The ‘reference inference module’ from SeqScreen-Nano (SeqScreen version 4.0) then used the filtered taxids from the Centrifuge to download the corresponding reference genomes from NCBI. All reads were then independently mapped using ‘minimap2’ to each of those individual reference genomes, and mapping metrics (e.g., depth, breadth of coverage) were calculated by SeqScreen-Nano. High-coverage reference genomes were concatenated by SeqScreen-Nano’s ‘reference inference module’ before the second alignment step, where the reads were remapped to the concatenated reference to allow each read to competitively map to the single best reference. Mapping metrics were re-calculated and further statistical analysis was performed to determine the likelihood of presence or absence of individual genomes based on a rubric. The SeqScreen-Nano ‘reference inference module’ analysis, metrics, and rubrics are originally described in Balaji et al. (2023a). Modification to the threshold criteria was made to be more inclusive of the reference genomes taken from the first mapping of individual reference genomes to the second mapping of a concatenated reference. This included reducing the original calculated in SeqScreen-Nano ‘coverage score’ threshold, which is the ratio of the observed ‘breadth of coverage’ and ‘expected breadth of coverage’ for each taxon in a sample (Balaji et al. 2023a), from a minimum of 0.70 to 0.10. The large ratio of 0.7 for the ‘coverage score’ was not ideal for smaller genomes for viruses, which had inflated expected coverages compared to genomes sizes of bacteria. This allowed for more reference viral genomes within the sample to be used in second stage mapping, while still removing potential contaminants. Additionally, signals from low abundance or partially covered genomes of close neighboring viral taxons were successfully reduced by requiring a greater than 0.15 ‘breadth of coverage’ for a reference genome to move forward in the second stage of mapping. These changes increased the inclusion of viral genomes known to be present within a sample while also reducing background and known contamination. The rubrics within SeqScreen v4.0 for “Present” (Species) and “Genus Present” calls were not modified.

### Evaluation of Untargeted Sequencing and Analysis

Statistical analysis and visualizations were performed using R version 4.3.1 (R Core Team 2023) and RStudio version 2023.06.01.524 (Posit team 2023). Visualization of the data was performed using the ‘ggplot2’ R package (Wickham 2016).

Limits of detection were determined for each of the seven representative RNA viruses in their respective clinical matrices by probit analysis using a series of dilutions across a minimum of 6-Log range (Table 4). The contrived samples in the plasma matrix were sequenced across a total of 8 multiplex runs and samples in VTM were sequenced across a total of 6 multiplex runs. Assay LoD were calculated in R and RStudio using probit regression model using the ‘glm’, generalized linear model, function as the concentration at which RNA virus was detected in 95% of replicates with at least 2 samples performed at each tested concentration (Table S3).

Precision was determined using repeat analysis of the contrived samples for RNA viruses at the load above and nearest their individual LoD as determined by probit analysis (LoD load) across 4 or 5 runs with VTM contrived samples and 4 or 7 runs with plasma contrived samples for inter-assay reproducibility. Precision was also determined using repeat analysis of NTC samples across all 14 runs for inter-assay reproducibility. Replicate remnant samples were also used for inter-assay reproducibility ranging from 2 to 3 independent runs performed for each sample performed across a total of 16 runs.

Precision for intra-assay reproducibility was determined for each individual RNA virus for duplicate contrived samples at LoD load performed on 4 to 7 runs. Intra-assay reproducibility was also determined for negative detection remnant samples representing 3 (plasma/serum) or all 6 (VTM) samples across a total of 2 runs.

Accuracy was determined using 24 remnant VTM samples comprised of 18 positive samples containing 6 detected organisms (three samples for each organism) and 6 negative samples. Sensitivity and specificity of the untargeted sequencing assay were calculated relative to prior ‘gold standard’ clinical molecular method, which was information provided with the obtained remnant samples.

To evaluate the effects of potential interfering substances, EDTA, human blood, and microbial contamination simulated by *Micrococcus luteus* or *Staphylococcus epidermidis* were added to contrived plasma samples. The addition of interfering substances was performed on plasma samples with RNA viral load level E (Table 4). EDTA was added to an additional 5 mM, 10 mM, and 100 mM EDTA disodium salt to the plasma samples prior to RNA extraction and downstream workflow. Blood was added to represent 2% or 5% of the total volume of the plasma sample. Additional analysis of the effects of human blood on the untargeted sequencing assay was performed on plasma samples with RNA virus loads of D, E, and F with the addition of 2% blood. For addition of *Micrococcus luteus* or *Staphylococcus epidermidis*, an overnight culture was washed twice with sterile molecular grade water. The cells were resuspended at high concentration and quantified by OD_600_. The bacteria were added to contrived plasma samples to generate bacterial concentrations at 10^7^ (high), 10^6^ (medium), and 10^5^ (low) cells per mL. The number of reads were determined by the Centrifuge results mapping to the individual RNA viruses. The read counts were normalized by each organism prior to performing ANCOVA in R to determine if significant differences existed between the concentration of interfering substances and no addition. Post-hoc comparisons between the concentration and no addition were made using TukeyHSD (honestly significant difference) testing.

An additional examination of remnant samples containing multiple viral detections or single sample representatives of a virus was performed. These 9 samples represented 10 different viruses and were sequenced using the untargeted sequencing assay and concordance with ‘gold standard’ clinical molecular testing was examined.

### Software availability

The “SigSciDx” computational software used in this study, along with the custom Centrifuge database, is publicly available as the following docker image on Docker Hub: sigsci/sigscidx (Signature Science Team 2023b). The version used for analysis in this publication is 1.0.1. A bash command to initiate the EPI2ME Labs framework is available on GitHub (Signature Science Team 2023c). Note that the database used for the custom viral Centrifuge database was from the June 2023 distribution of GenBank genome assemblies.

## Data Access

Metagenomic reads from contrived and remnant samples from this study were depleted of human host sequences and will be submitted to the NCBI BioProject database (Signature Science Team 2023a). The MS2 Phage genome sequence used was NC001417.2 for mapping. The *gfp* gene sequence used for mapping was obtained through direct communication with the vendor, System Biosciences, LLC (MR700A-1). Both MS2 Phage and *gfp* gene sequences from the specific vendors are included in the SigSciDx docker image.

## Data Availability

All data produced in the present study are available upon reasonable request to the authors. Metagenomic reads from samples from this study will be depleted of human host sequences and will be submitted to the NCBI BioProject database before peer reviewed publication.

## Acknowledgements

The authors would like to thank Leslie Parke for oversight and project management for this effort. The authors would also like to thank Jim Gibson for his assistance creating the workflow figure. The authors would also like to thank Leslie Parke for her review. This work was supported by the Centers for Disease Control and Prevention under contract number 75D30122C15359.

## Authors’ contributions

All authors have read and approved the manuscript. Conceptualization: ADK, KQS, MBS, FCH. Data curation: ADK, MBS, NCK. Laboratory analysis: ANS, NCK, KQS, EAS, CJS, ADK, LWA, TAW. Funding acquisition: ADK, FCH, KLT, KQS. Data analysis: ADK, MBS, NCK, KLT, FCH. Project administration: ADK, FCH. Writing – original draft: ADK, NCK, MBS, KQS, VMJ, FCH, KLT. Writing – review & editing: ADK, KQS, MBS, LWA, KLT, FCH.

---

**Table S1.**
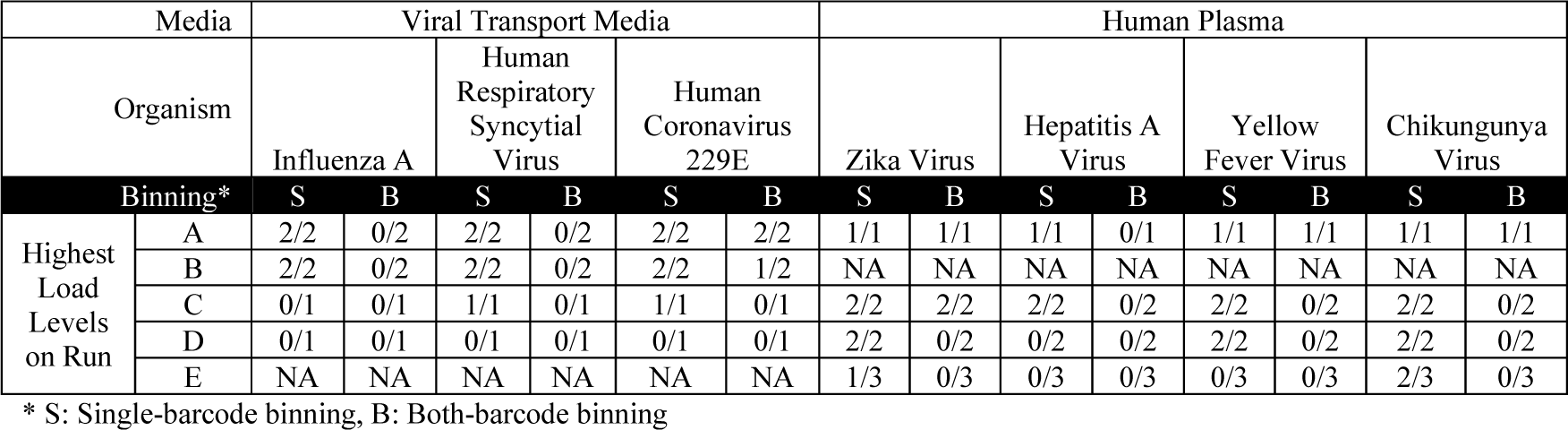
Indication of Barcode Crosstalk by Detection of RNA Virus in TNC.

| Media |  | Viral Transport Media |  |  |  |  |  | Human Plasma |  |  |  |  |  |  |  |
| --- | --- | --- | --- | --- | --- | --- | --- | --- | --- | --- | --- | --- | --- | --- | --- |
| Organism | Binning* | Influenza A |  | Human Respiratory Syncytial Virus |  | Human Coronavirus 229E |  | Zika Virus |  | Hepatitis A Virus |  | Yellow Fever Virus |  | Chikungunya Virus |  |
|  |  | S | B | S | B | S | B | S | B | S | B | S | B | S | B |
| Highest Load Levels on Run | A | 2/2 | 0/2 | 2/2 | 0/2 | 2/2 | 2/2 | 1/1 | 1/1 | 1/1 | 0/1 | 1/1 | 1/1 | 1/1 | 1/1 |
|  | B | 2/2 | 0/2 | 2/2 | 0/2 | 2/2 | 1/2 | NA | NA | NA | NA | NA | NA | NA | NA |
|  | C | 0/1 | 0/1 | 1/1 | 0/1 | 1/1 | 0/1 | 2/2 | 2/2 | 2/2 | 0/2 | 2/2 | 0/2 | 2/2 | 0/2 |
|  | D | 0/1 | 0/1 | 0/1 | 0/1 | 0/1 | 0/1 | 2/2 | 0/2 | 0/2 | 0/2 | 2/2 | 0/2 | 2/2 | 0/2 |
|  | E | NA | NA | NA | NA | NA | NA | 1/3 | 0/3 | 0/3 | 0/3 | 0/3 | 0/3 | 2/3 | 0/3 |

\* S: Single-barcode binning, B: Both-barcode binning

**Table S2.** Single-detection remnant sample calls.

| Sample | Original Detection Method | Untargeted Sequencing Call |
| --- | --- | --- |
| Human Metapneumovirus | Genmark Eplex PCR | Not Detected, Internal Control (IC) Quality Control (QC) failed |
| Human Metapneumovirus | Genmark Eplex PCR | Not Detected |
| Human Metapneumovirus | Genmark Eplex PCR | Human metapneumovirus, IC QC failed |
| Parainfluenza IV | Genmark Eplex PCR | Not Detected |
| Parainfluenza IV | Genmark Eplex PCR | Human parainfluenza virus 4a |
| Parainfluenza IV | Genmark Eplex PCR | Not Detected |
| SARS-CoV-2 | Gene Xpert Infinity | Severe acute respiratory syndrome coronavirus 2 |
| SARS-CoV-2 | Gene Xpert Infinity | Severe acute respiratory syndrome coronavirus 2 |
| SARS-CoV-2 | Gene Xpert Infinity | Severe acute respiratory syndrome coronavirus 2 |
| RSV | Biofire | Respiratory syncytial virus |
| RSV | Biofire | Respiratory syncytial virus |
| RSV | Biofire | Respiratory syncytial virus |
| Influenza A | Biofire | Influenza A |
| Influenza A | Diasorin Integrated Cyclor | Influenza A |
| Influenza A | Biofire | Influenza A |
| Enterovirus | Diasorin Integrated Cyclor | Rhinovirus A1 |
| Enterovirus | Diasorin Integrated Cyclor | Rhinovirus A1 |
| Enterovirus | Diasorin Integrated Cyclor | Rhinovirus A1 |

**Table S3.** Number of contrived samples at virus load sequenced.

| Load Level<br>(See Table 5) | Plasma | VTM |
| --- | --- | --- |
| A | 2 | 4 |
| B | 2 | 8 |
| C | 6 | 10 |
| D | 8 | 8 |
| E | 14 | 4 |
| F | 10 | 2 |
| G | 6 | NA |

